# Auditory Enhancement of Sleep Slow Waves in People with Parkinson’s Disease: A Proof-of-Concept Study

**DOI:** 10.1101/2025.01.30.25320306

**Authors:** Simon J. Schreiner, Jana Horlacher, Sara Fattinger, Klavs Renerts, Luzius Brogli, Lydia Kämpf, Maurizio Scandella, Rositsa Poryazova, Philipp O. Valko, Laura Ferster, Renske Sassenburg, Valeria Jaramillo, Giulia Da Poian, Caroline Lustenberger, Walter Karlen, Reto Huber, Christian R. Baumann, Angelina Maric

## Abstract

Deep sleep supports several restorative functions and has emerged as a potential therapeutic target in neurodegenerative disorders, such as Parkinson’s disease (PD). Phase-targeted auditory stimulation (PTAS) is a non-invasive method for enhancing slow waves of deep sleep, measurable by increased slow-wave activity (SWA; spectral power 0.75–4.5 Hz). Here, we report the first study investigating whether PTAS can be successfully applied in PD patients with subjective sleep disturbance.

We conducted a randomized, double-blind, sham-controlled, cross-over, within-subject study assessing the effects of PTAS over three nights with wearable devices at participants’ homes. PTAS was applied in ON and OFF windows for the entire night in 14 participants and first part of the night in nine.

PTAS increased low-frequency SWA by almost 30% during ON windows in 21 analyzed participants. The SWA increased further with higher numbers of stimuli, suggesting a dose-dependent effect. Participants receiving part-night PTAS reported improved subjective daytime sleepiness after the third night.

This proof-of-concept study demonstrates successful auditory enhancement of slow waves with PTAS in PD. Our observation of the potential advantages of part-night PTAS warrant further exploration. Our data encourage longer and larger trials to fully explore the potential of PTAS in PD and other neurodegenerative diseases.

## Introduction

Parkinson’s disease (PD) is the second most common neurodegenerative disease after Alzheimer’s disease (AD). Although 1–2% of older adults aged 65 and above already have PD^1^, its incidence and related economic burden are expected to further rise exponentially^2,3^. PD is slowly progressive and currently incurable, often resulting in complications and loss of independence. Although PD is well-known for its cardinal motor symptoms, such as bradykinesia and tremor, nonmotor symptoms are key components of PD too^4^. Sleep–wake disturbances are among the most common and bothersome nonmotor symptoms of PD, with a significant impact on patients’ quality of life^4^. Sleep maintenance insomnia affects up to 90% of patients, excessive daytime sleepiness around 30%, and rapid eye movement (REM) sleep behavior disorder (RBD) between 30% and 80%, depending on age^5^. However, treatment options for nonmotor symptoms, especially sleep–wake disturbances, are scarce, and disease-modifying therapies remain a major unmet need.

Growing evidence supports a bidirectional relationship between sleep and neurodegeneration: Neurodegeneration causes sleep–wake disturbances, and disturbed sleep may in turn accelerate neurodegeneration^6^. Consequently, interventions that improve sleep offer unique opportunities for targeting neurodegeneration^7^. Deep sleep has emerged as an especially promising therapeutic target because several related functions may exert beneficial effects on neurodegeneration^8–10^. Deep sleep is defined by electroencephalographic (EEG) slow waves, which are large-amplitude oscillations in the delta frequency range (0.75–4.5 Hz) that result from synchronous neuronal firing in thalamocortical networks^11^. In PD, translational evidence suggests that deep sleep worsens progressively from preclinical to late stages^12–16^. Deep-sleep deficiency in PD correlates with daytime sleepiness^17^, disease milestones such as dyskinesia and cognitive impairment^18–20^, putative imaging markers of altered brain clearance^21^, and clinical progression^22–25^. Importantly, enhancing deep sleep pharmacologically alleviates sleep– wake disturbances in PD patients^26^, and even reduces alpha-synuclein accumulation, the pathological hallmark of PD, in mice^27^. Mechanistically, sleep could regulate accumulation of alpha-synuclein and other pathological proteins, such as ß-amyloid, by reducing their production or increasing clearance^28–30^. Critically, modulation of deep sleep, rather than sleep per se, influences pathological proteins^31,32^. Pharmacological enhancement of deep sleep is limited by side effects and tolerance issues and may alter sleep macrostructure. Thus, nonpharmacological methods that are well tolerated and allow targeted modulation of specific sleep oscillations are particularly promising, especially in older clinical populations ^8,9,26,33^.

Phase-targeted auditory stimulation (PTAS) is a nonpharmacological method of modulating slow waves, the EEG hallmark of deep sleep. Real-time or closed-loop acoustic stimulation are alternative names for PTAS. This method works by presenting non-arousing acoustic stimuli synchronized to a specific phase of endogenous slow waves^34^. PTAS has been shown to enhance slow wave amplitudes when targeting the up-phase of slow waves, which is reflected in increased EEG spectral power in the corresponding frequency range, slow wave activity (SWA, 0.75–4.5 Hz)^34–36^. Importantly, enhancing slow waves with PTAS has been shown to benefit deep-sleep-related functions, such as overnight memory consolidation^34^, immunological processes^37^, and even levels of pathological proteins^38,39^. Novel wearable devices enable self-applied PTAS over multiple nights at home, which is an important prerequisite for translating enhancement of slow waves with PTAS into a clinical application for therapeutic purposes^40–42^. However, PTAS is less efficient in healthy older adults and those with mild cognitive impairment than in healthy young adults^41,43–45^. Moreover, it is unknown whether PTAS can be used to enhance slow waves in people with PD, who in addition to older age show complex sleep–wake disturbances and altered sleep physiological processes^5,46,47^. In particular, PD patients show deep-sleep deficiency compared to controls, with reduced SWA, lower slow wave density, and shorter deep-sleep duration^14,48,49^.

In the current study, we translated a novel approach for self-applied, at-home PTAS, recently successfully performed in healthy older adults^41^, into an application in PD patients with subjectively disturbed sleep (Figure 1). Translation included the personalization and optimization of sleep detection and PTAS algorithm settings to account for the high variability in sleep EEG characteristics in this patient population. We aimed to evaluate the potential of PTAS to enhance slow waves and to investigate possible effects of PTAS on sleep macrostructure and the subjective sleep–wake experience.

**Figure 1:**
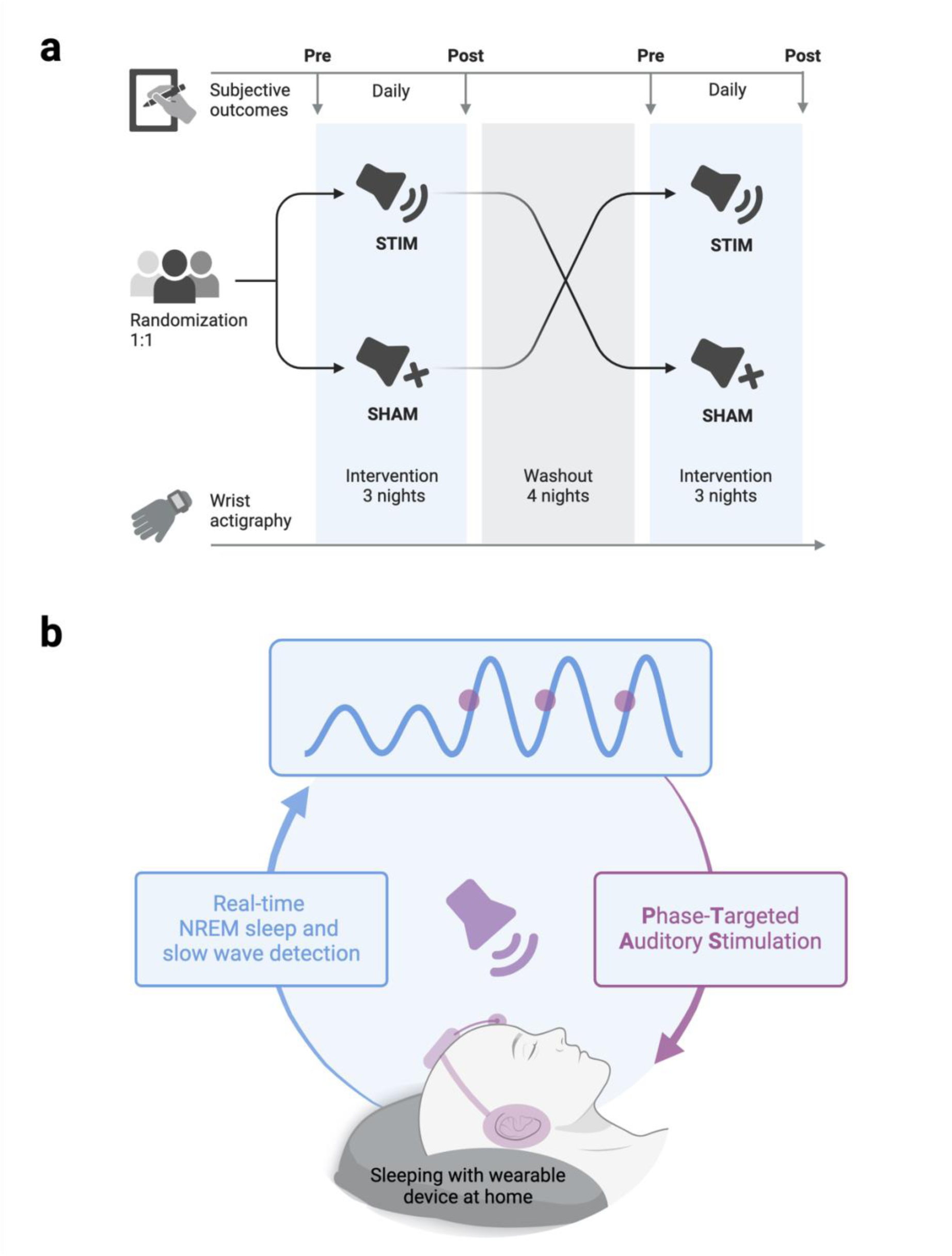
Study design a) This is a randomized, counterbalanced, double-blind, sham-controlled, crossover study comparing the effects of three nights of phase-targeted auditory stimulation (PTAS) with three nights of sham stimulation on sleep in people with Parkinson’s disease and subjective sleep disturbance. b) PTAS is a nonpharmacological method for enhancing slow waves, the EEG hallmark of deep sleep. This non-invasive method works through administration of brief (50 ms) nonarousing stimuli of multifrequency pink noise that precisely target the up-phase of endogenous slow waves. Participants used the Mobile Health Systems Lab SleepBand v3 (MHSL-SB), a wearable device for at-home sleep recording and PTAS, based on real-time EEG processing and automated detection of sleep and presence of slow waves40,41. During one intervention period, the device applied PTAS using a windowed ON– OFF approach (STIM). During the other intervention period, the muted device recorded EEG along with potential stimuli (SHAM). Outcomes included sleep EEG features recorded by the MHSL-SB and subjective sleep–wake experience assessed by daily questionnaires throughout the interventions. Created with biorender.com.

We hypothesized that the effects of PTAS observed in previous studies, with increased SWA as a main effect, can be replicated in PD patients when PTAS settings are tailored to the deep-sleep alterations seen in this clinical population. We further hypothesized that PTAS may improve subjective outcomes of sleep–wake experience.

## Results

Figure 2 shows a patient flowchart, and Table 1 summarizes patient characteristics. We randomized a total of 23 participants. After completing the study in the first 14 participants, we adapted the PTAS protocol to restrict PTAS to the first part of the night for the following nine participants in the STIM condition (see Methods and Discussion for more details), resulting in two subgroups (Table S1). Two individuals had to be excluded from analysis, resulting in a total of 21 participants in the final analysis, of which 13 had received whole-night and eight part-night PTAS. The absence of PTAS-induced increase in SWA during screening was an exclusion criterion that applied to five out of 33 participants (15 %) that had undergone a screening night.

**Figure 2:**
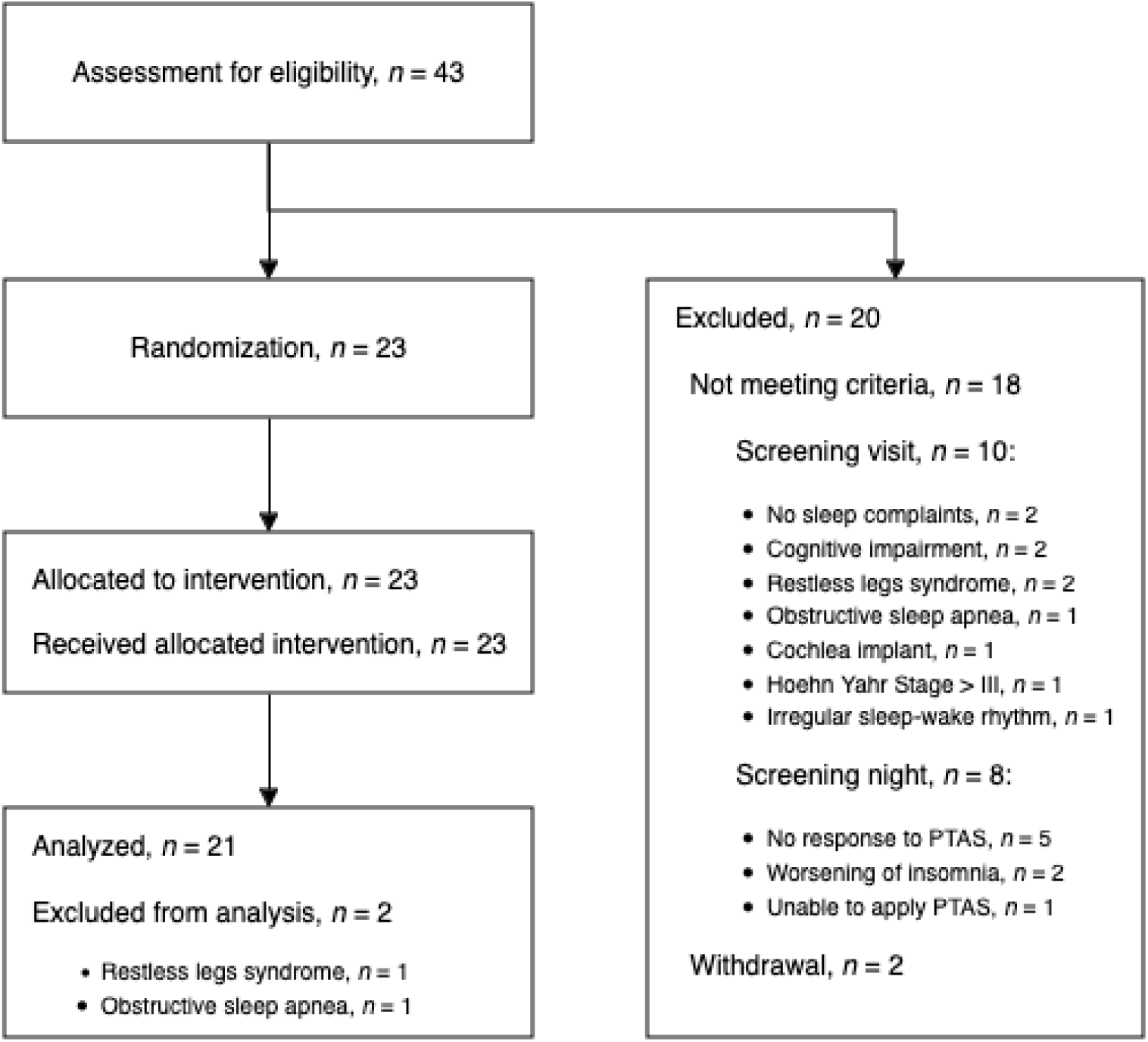
Patient flow chart Of 43 subjects who underwent screening, 23 were randomized, 18 excluded due to not meeting the criteria, and two decided against participation after successful screening. See Methods section for more details.

**Table 1:** Patient characteristics.

| Characteristics | *n* = 21 |
| --- | --- |
| Age (years) | 58 (53, 60) |
| Females | 10 (48%) |
| Disease duration (years) | 5 (4, 7) |
| Hoehn & Yahr Stage |  |
| II | 20 (95%) |
| III | 1 (4.8%) |
| MDS-UPDRS III | 22 (17, 30) |
| Levodopa equivalent dose (mg/d) | 650 (510, 831) |
| Body Mass Index (kg/m<sup>2</sup>) | 26.1 (24.3, 27.1) |
| Parkinson's Disease Sleep Scale (PDSS-2) | 17 (15, 25) |
| PDSS-2 disturbed sleep subscale | 9 (8, 11) |
| Epworth Sleepiness Scale | 11 (8, 14) |

Table shows median values (interquartile range).

### Slow-wave activity

To analyze the immediate effects of PTAS on non-rapid eye movement (NREM) sleep EEG power, we analyzed the common stimulation time in all participants, which is the first part of the night. We found significant differences in NREM sleep EEG power between STIM and SHAM, with an increase in the low-frequency range of SWA (1–1.25 Hz) similar to that observed in healthy older adults^41^. We also found a decrease in the adjacent mid-frequency range of SWA (1.5–2.25 Hz) (Figure 3 a). Given these mixed differences in SWA when analyzing all NREM sleep windows, we next compared EEG power between STIM and SHAM for ON windows, during which stimulation was applied, and for subsequent OFF windows, when it was not, separately. Here, we found that PTAS increased SWA by up to 29.52 ± 16.82 % between 0.75 Hz and 1.5 Hz during ON windows (Figure 3 b). Conversely, SWA decreased by up to 11.85 ± 6.48 % between 0.75 Hz and 3.5 Hz and at 4.5 Hz during subsequent OFF windows (Figure 3 b). Therefore, the mixed differences in SWA across all NREM sleep windows corresponded to the ON–OFF-related dynamics of SWA modulation, with strong increases in the low-frequency range when stimulation was ON and mild decreases in the entire SWA frequency range after stimulation when stimulation was OFF. This pattern of SWA redistribution between ON and OFF windows is in line with previous PTAS studies in older adults and patients with mild cognitive impairment^43,44^.

**Figure 3:**
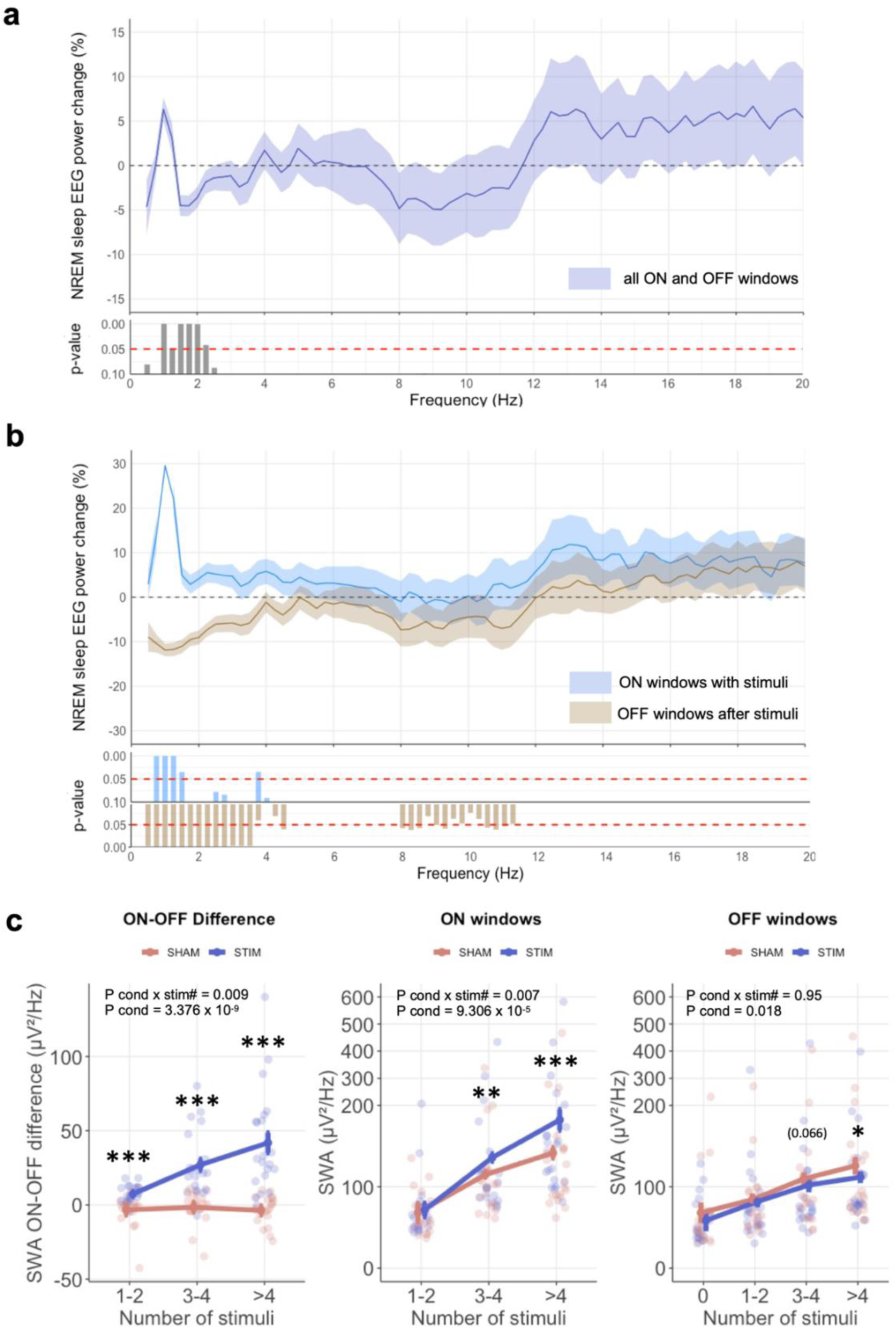
Effects of phase-targeted auditory stimulation (PTAS) on NREM sleep EEG power **a:** The y-axis displays relative differences between STIM and SHAM in normalized EEG power across frequency bins between 0.5 Hz and 20 Hz for all windows of NREM sleep during the first part of the night, the common stimulation time across all participants. The *p*-values above the dotted red line indicate *p* <0.05 (robust linear mixed model (RLMM), fixed factor condition) within each frequency bin (see methods). **b:** The y-axis displays the equivalent differences in relative NREM sleep EEG power as in Figure 3 a but for ON windows with at least one stimulus (light blue color) and subsequent OFF windows (beige color) separately. Note that the values are relative to the corresponding ON and OFF windows during SHAM. The *p*-values above or below the dotted lines indicate *p* <0.05 (RLMM, fixed factor condition) within each frequency bin (see methods). **c:** Plots show the dose-dependent relationship between number of stimuli (stim#) and changes in slow wave activity (SWA) 0.75–4.5 Hz for differences between ON and OFF windows (left), mean SWA during ON windows (middle), and mean SWA during OFF windows after stimulation (right). The increasing levels of SWA along the number of stimuli during ON and OFF windows in SHAM indicate that more stimuli can be applied when more slow waves are present and SWA is higher as a consequence. The effect of the intervention is seen in the difference between the STIM and SHAM trajectories. In the left and middle plots, values are categorized and matched for the number of stimuli during STIM (blue color) or potential stimuli during SHAM (red color). In the right plot, values are categorized and matched for the number of stimuli during preceding ON windows. Asterisks indicate *p* <0.05 (*), *p* <0.01 (**), and *p* <0.001 (***) from the RLMM analysis within each category of number of stimuli. Parentheses indicate results from exploratory analyses that followed an insignificant interaction. All N = 21. Note that y-axes are condensed for illustrative purposes.

Another study in healthy older adults had suggested a dose-dependent effect of PTAS, with more stimuli applied during ON windows leading to greater SWA increases^41^. We confirmed this dose dependence with a robust linear mixed model (RLMM) analysis that showed a significant interaction between the condition (STIM vs. SHAM) and number of stimuli (#nstim) factors (see Methods section Statistical analysis) on SWA (Figure 3 c, left and middle). The RLMM suggested that the decrease in SWA in subsequent OFF windows was not dose dependent (Figure 3 c, right). However, exploratory analyses suggested that differences between STIM and SHAM in SWA during OFF windows were only significant when the preceding ON windows contained four or more stimuli (Figure 3 c, right).

### Sleep macrostructure

We found no effects of PTAS on sleep macrostructure when analyzing the first part of the night, the common stimulation time in all participants (Table S2). We then ran separate whole-night analyses on participants who had received PTAS for the entire night (whole-night subgroup, *n* = 13 for analysis) or only the first part of the night (part-night subgroup, *n* = 8 for analysis). Here, we observed trends towards more whole-night wake after sleep onset (WASO; RLMM: F condition = 4.55 (1,8.9), *p* = 0.062) and lower whole-night sleep efficiency (SE; RLMM: F condition = 5.11 (1,8.9), *p* = 0.05) during STIM than SHAM, but only in the whole-night subgroup (Table S3). This trend was evident for the time spent in WASO (min) but not for the number of awakenings (RLMM: F condition = 0.71 (1,9.6), *p* = 0.419), indicating a tendency for longer WASO but not more frequent awakenings during STIM than SHAM in participants who had received PTAS for the entire night. Notably, this trend was not evident when analyzing only the first part of the night in these participants (results not shown), which may indicate potential disadvantages for sleep continuity when PTAS is applied for the entire night. In line with this assumption, we observed no effects of STIM compared to SHAM on whole-night sleep macrostructure in participants receiving PTAS only during the first part of the night (Table S3).

### Subjective outcomes

We tested the effects of PTAS on subjective outcomes, including sleep quality, daytime sleepiness, and mood, assessed by daily visual analogue scales (VAS), daily momentary sleepiness assessed with the Karolinska Sleepiness Scale (KSS), and PD-related sleep disturbances assessed with the 15-item Parkinson’s Disease Sleep Scale, version 2 (PDSS-2) and its disturbed sleep subscale pre- and postintervention^50^. Previous studies suggested that any beneficial effects of PTAS might require several nights of intervention^38,39^. Therefore, we also tested for interaction effects between the condition and night factors. We found a significant interaction of condition and night on subjective daytime sleepiness assessed by VAS (F condition x night = 6.95 (1,24.55), *p* = 0.014). Investigating this effect further, we found that PTAS was associated with a reduction in daytime sleepiness, but only after the third and last nights and not after the first two nights, and only in the part-night subgroup but not the whole-night subgroup (Figure 4).

**Figure 4:**
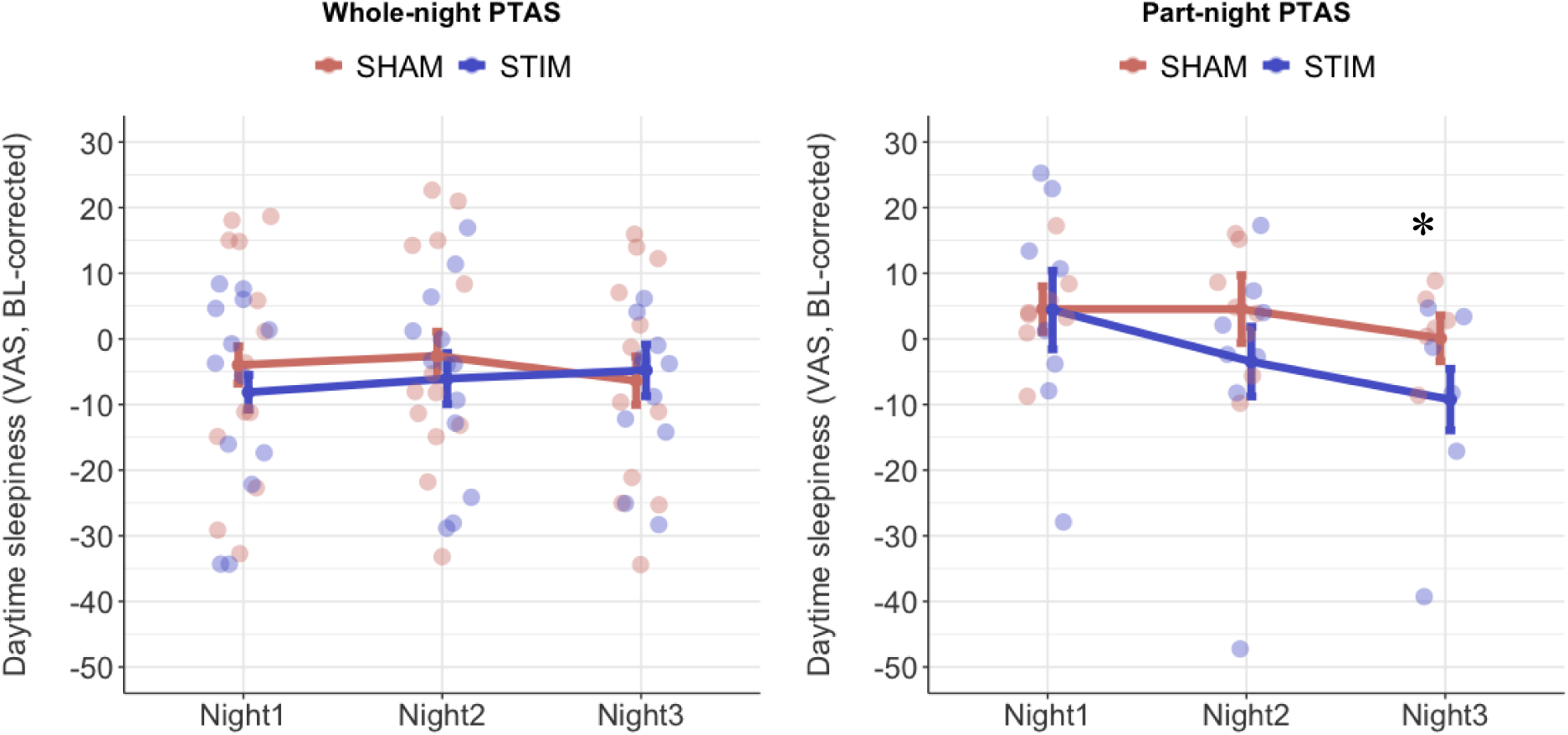
Effects of phase-targeted auditory stimulation (PTAS) on subjective daytime sleepiness The Y-axis shows changes from baseline (BL, means, and SEM bars) in VAS ratings (0–100) of daytime sleepiness across the three intervention nights in the two subgroups that received whole-night PTAS (left, *n* = 13) or part-night PTAS (right, *n* = 8). Only in the part-night subgroup was the interaction of condition (STIM, SHAM) and night (Night 1, Night 2, Night 3) significant (robust linear mixed model (RLMM): F condition x night = 6.95 (1,24.55), *p* = 0.014), with significant differences after Night 3. Asterisk (*) indicates *p* <0.05 resulting from RLMM analyses performed for each night separately.

We did not observe other changes in subjective ratings as a function of condition or its interaction with night (Table S4).

Because daytime sleepiness did not improve and sleep continuity (longer WASO, lower SE) showed worsening trends in the whole-night subgroup, we assessed whether changes in sleep continuity due to whole-night PTAS may have negatively influenced daytime ratings. We performed exploratory correlation analyses between changes in WASO and SE and differences in KSS and VAS ratings on sleep quality, daytime sleepiness, and mood. We averaged daily measures of WASO, SE, and subjective outcomes during each intervention and calculated relative differences in WASO and SE and differences in KSS and VAS ratings between STIM and SHAM. This analysis revealed no associations of differences in WASO or SE with differences in the KSS or VAS ratings (all *p* >0.47).

## Discussion

This study is the first to establish proof of the concept that phase-targeted auditory stimulation (PTAS) can enhance slow waves, the EEG hallmark of deep sleep, in Parkinson’s disease (PD) patients with subjectively disturbed sleep. We administered PTAS over three nights using a wearable device at home, for the entire night in 14 and the first part of the night in nine participants. Our main finding is that PTAS increases slow-wave activity (SWA) in PD patients, in a dose-dependent manner. We further found that part-night PTAS improved subjective daytime sleepiness without significantly influencing sleep macrostructure. These effects were not observed with whole-night PTAS. Overall, these encouraging findings underscore the potential of PTAS as a non-invasive, nonpharmacological neurostimulation method for boosting deep sleep and related functions, even in clinical populations with neurodegenerative diseases and disturbed sleep such as PD.

### Effects of PTAS on sleep in PD

Although the present study population largely differs from the mostly healthy participants in previous studies, we show that the effects of PTAS on SWA in such a clinical population are comparable to the effects in previous studies.

First, we showed that PTAS increases low-frequency SWA during NREM sleep. Interestingly, PTAS showed mixed effects within the SWA frequency range during and immediately after stimulation: whereas SWA in the low frequency range increased strongly when stimulations were applied during ON windows (around +30% in our study), SWA in the entire delta frequency range decreased mildly after stimulations during subsequent OFF windows (around -10% in our study). This pattern resembles previous observations on SWA redistribution or reorganization, between ON and OFF windows described in healthy older adults and patients with mild cognitive impairment^43,44^. The relevance of the PTAS-induced SWA redistribution remains unclear, but it could reflect a positive reorganization of SWA: First, Papalambros et al. reported that the degree of SWA redistribution between ON and OFF windows related to improvements in overnight memory consolidation^43,44^. Second, Winer et al. reported strikingly different effects of low and high-frequency SWA on AD pathology, with SWA <1 Hz relating to less and SWA >1 Hz to more future accumulation of ß-amyloid in older adults^51^. Accordingly, the potential of PTAS for increasing SWA in the low frequency range has been proposed as a promising therapeutic approach to neurodegeneration^52^. Future studies should further explore the relevance of PTAS-induced changes in SWA and how SWA needs to be modulated exactly to promote deep sleep-related functions with beneficial effects in a context of neurodegeneration.

Other studies reported variable, strongly diminished responsiveness to PTAS in older individuals than younger ones without SWA redistribution^41,45^. It is worth mentioning that our study differs from these studies in that we excluded SWA nonresponders. This limits direct comparisons with prior studies that also included nonresponders. Notably, we only identified 15% as nonresponders during screening. This rate is low and highlights the successful translation of the PTAS approach to a clinical application.

Second, we confirmed that PTAS may increase SWA in a dose-dependent manner^41^. We found that higher numbers of stimuli lead to greater increases in SWA during ON windows. Interestingly, the decrease in SWA during OFF windows did not show a similar dose dependence. This observation suggests that the decrease in SWA during OFF windows is not directly proportional to the intensity of the preceding stimulation. However, exploratory analyses showed that the OFF decrease reached significance only after maximum amounts of stimuli during preceding ON windows.

We found new, albeit indirect, evidence that PTAS applied only for the first 2–3 hours of the night may have advantages over whole-night PTAS. Most evidence to date consistently shows that PTAS modulates microstructural features of NREM sleep, such as SWA, but does not impact sleep macrostructure^53,33^. However, a recent study in older adults showed tendencies for reductions in REM sleep, which correlated with worsening mood experience during PTAS compared to the sham condition^41^. A reduction in REM sleep due to PTAS may appear to be a plausible explanation for such an effect on mood, especially when PTAS is applied during the second half of the night, when REM sleep and low-amplitude NREM sleep predominate. It is conceivable that increasing SWA during late NREM sleep alters the dynamics between NREM and REM sleep. If so, approaches that respect the temporal dominance of NREM and REM sleep may seem a beneficial choice, particularly for patients with neurodegenerative diseases who already show altered and often impaired REM sleep^48,54^. Therefore, after completing the study in the first 14 participants, we decided to restrict PTAS to the first part of the night, during which deep sleep physiologically predominates, for the following nine participants. Our aim was to achieve effects of PTAS on NREM sleep while avoiding potential impact on REM sleep or overnight dynamics of SWA with physiologically low levels of SWA during late sleep. Interestingly, we observed that the intervention led to decreased sleep continuity when PTAS was applied for the entire night. In contrast, we observed no effects on sleep macrostructure when we applied PTAS only for the first part of the night. However, we did not observe any effects of PTAS on REM sleep, regardless of how long PTAS was applied during the night. One explanation for this difference from previous findings on the impact of whole-night PTAS on REM sleep in healthy older adults is that the amount of REM sleep is reduced in patients with neurodegenerative diseases, such as PD^48^. Importantly, we did not find evidence that the subtle changes in sleep macrostructure during whole-night PTAS had an impact on subjective daytime ratings. Moreover, it should be stressed that our findings indicated longer wake time after falling asleep but not more frequent awakenings during STIM compared to SHAM nights. One possible explanation is that potential awakenings during late NREM sleep might be associated with difficulties falling asleep again, especially in a clinical population with sleep disturbance. Moreover, it is possible that the changes in sleep macrostructure during STIM were not primarily related to PTAS but to other reasons uncontrolled by the experimental design, for example, fluctuations in chronic sleep disturbance.

However, the current study was not designed to systematically compare protocols with different overnight durations of PTAS, because we changed our protocol halfway through the study due to new evidence at that time^41^. Consequently, participants who received the protocols with different PTAS durations were not matched in demographic and clinical characteristics, which prevents direct comparisons and definitive conclusions on this matter (Table S1). For example, the proportion of females was higher among participants with whole-night PTAS than among those with part-night PTAS. Given that sex differences exist in sleep and neurodegenerative disease, our observations could also be driven by variable responses to PTAS as a function of sex. Future studies should compare the effects of whole-night PTAS with part-night PTAS more systematically, because aligning PTAS to the homeostatic overnight dynamics of SWA could have certain advantages.

### Clinical effects of PTAS in PD

We found evidence that PTAS potentially provides delayed clinical benefits. Specifically, we observed a delayed improvement in daytime sleepiness due to PTAS, which became detectable after two nights and statistically significant after three nights of PTAS in the part-night subgroup. Excessive daytime sleepiness in PD is multifactorial and largely driven by PD pathology because it is independent from nocturnal sleep according to most studies ^55^. However, recent evidence connects daytime sleepiness in PD to deep-sleep deficiency at the microstructural level^17^. Our observation of delayed improvement in daytime sleepiness is reminiscent of the delayed improvement in daytime sleepiness in patients with PD or narcolepsy treated with pharmacological slow-wave enhancement, which only fully sets in after several weeks of treatment^26,56^. Therefore, our study adds more evidence suggesting that slow-wave enhancement is s potential strategy for treating daytime sleepiness in PD. Nevertheless, we consider this finding preliminary because of the small sample size in the part-night PTAS subgroup. Moreover, we did not observe improvements in other subjective outcomes. Furthermore, clinical improvement in the part-night subgroup but not the whole-night subgroup could indicate potential advantages of part-night PTAS in our sample.

To the best of our knowledge, previous PTAS studies did not include clinical populations with sleep disturbances and did not systematically assess the effects of PTAS on subjective sleep– wake experience. However, previous work has suggested that PTAS may have delayed effects on clinical outcomes, as shown in the improvement of memory performance in older adults^38,39^. Therefore, it is conceivable that prolonged administration of PTAS could be necessary and that the intervention of three nights applied in the present study was too short to elicit more robust clinical benefits. Accordingly, longer clinical trials applying PTAS over several weeks or months might be needed to fully assess its clinical efficacy. The availability of wearable devices for at-home PTAS offers exciting opportunities for exploring its long-term effects in humans.

## Limitations

Our study has several limitations. First, due to the remote study design, it was impossible to control for the potential impact of several factors that may have influenced the outcome of this study, even with the chosen within-patient design. These factors include all aspects of behavior that can impact on sleep, including various kinds of daytime activities such as sports and napping, light exposure, bedtime routines, and dietary factors. Consequently, study participants were instructed to adhere to their regular routines as closely as possible, and for example, subjects with highly irregular sleep–wake patterns were excluded from the study. Nevertheless, a certain degree of freedom is an unavoidable part of any study that aims at translating PTAS from controlled single-night in-lab experiments to real-world at-home applications.

As mentioned above, we adjusted our protocol in the overnight duration of PTAS, without previously intending to compare PTAS protocols. This may have reduced statistical power for detecting certain effects of PTAS on clinical outcomes. Although we were able to pool the data for the immediate stimulation effect on NREM sleep EEG by restricting the analysis to the common stimulation time, next-day ratings are always influenced by the whole sleep period, resulting in different conditions for the two subgroups. Our observations on potential advantages to restricting PTAS to the deep-sleep-rich first part of the night are certainly of interest but need to be considered as anecdotal at this point.

We included patients with subjective sleep disturbances as part of the study design, but objective sleep assessment, for example with polysomnography, was not included in the screening. Therefore, the risk of unknown objective sleep disturbances, such as covert sleep apnea, is possibly higher among our study sample than among participants identifying as good sleepers in other studies. Nevertheless, the recruitment via our outpatient clinic and neurological practices as well as the thorough history taking and continuous sleep EEG monitoring during screening and throughout the study provided additional information. Consequently, participants could be preselected for the presence of known sleep apnea or restless legs syndrome, and we excluded five participants diagnosed with these conditions, three during screening, and two after completing the study. Symptoms of restless legs syndrome are frequent in PD patients, fluctuate naturally, and are difficult to distinguish from other nocturnal symptoms, especially nocturnal wearing off, i.e. the worsening of motor symptoms when the effect of dopaminergic medication wears off during the night. Therefore, depending on their intention, future studies may consider polysomnography or combination with home screening for sleep apnea or periodic limb movements during sleep as regular elements of the screening process, or at least keep a low threshold for additive testing in suspected cases.

Because we aimed to assess the potential of PTAS in PD to enhance SWA, we excluded five out of 33 patients who showed no response to PTAS during the screening nights. Therefore, our study should not be used to judge the efficacy of PTAS in PD patients. The resulting rate of 15% nonresponders is low in comparison with the range of responses in previous studies in older adults and a recent report in people with AD^57^. Notably, we may have optimized the rate of responders by individual adaptations of volume and algorithm thresholds for NREM sleep and slow wave detection, which could optimize the application of PTAS in populations with low-amplitude slow waves, such as older adults and patients with neurodegenerative disease^58,59^. We excluded patients with deep brain stimulation; however, implanted deep brain stimulation devices show synergistic potential with PTAS applications^60^.

### Outlook and perspectives

The present study contributes to translational research on the therapeutic potential of sleep interventions in neurodegenerative disease. Evidence supporting a neuroprotective role of deep sleep in PD and other neurodegenerative diseases is growing, and our study encourages further applications of PTAS to explore the potential of nonpharmacological slow-wave enhancement for targeting neurodegeneration in greater detail. Some aspects of our study may extend to clinical populations with chronic sleep–wake disturbances due to other causes and encourage further pilot studies on this behalf.

Furthermore, we have demonstrated the successful application of a wearable device for at-home PTAS in a clinical population. Through remote monitoring and adaptation of settings, such wearable systems may help tailor PTAS to the specific needs of some clinical populations, for example, those characterized by low-amplitude slow waves or high variability in sleep EEG characteristics. Indeed, personalized sweet-spot appraoches could be a prerequisite for translating sleep modulation, such as PTAS, into large-scale clinical applications, especially in patients with altered sleep–wake functioning.

## Conclusion

This proof-of-concept study demonstrates the potential of PTAS as a nonpharmacological method for boosting sleep slow waves, the EEG hallmark of deep sleep, in people with neurodegenerative diseases such as PD. Future studies should apply PTAS over longer periods to fully explore its therapeutic potential for symptomatic benefits and disease-modifying properties.

## Methods

### Ethics

This study was conducted according to the Declaration of Helsinki principles and approved by the Cantonal Ethics Committee of Zurich (reference: BASEC 2020-00431) and Swissmedic, the Swiss agency for drugs and therapeutic products (reference: 10000681), and registered at ClinicalTrials.gov (NCT04589182). All participants gave written informed consent prior to inclusion in the study. Participants received 200 Swiss francs to reimburse expenses (e.g. travel to the study site).

### Study design and intervention

This was a single-center, 1:1 randomized, counterbalanced, double-blind, sham-controlled, crossover study conducted at the Department of Neurology, University Hospital Zurich, Switzerland. The study took place between Oct 2020 and May 2022, (whole-night subgroup: Oct 2020–Jul 2021; part-night subgroup: Sep 2021–May 2022). Due to the SARS-CoV-2 pandemic, the study design minimized personal contacts and visits to the study site and embraced remote assessments whenever possible to adhere to regulatory restrictions in pandemic times.

Figure 1 summarizes the study design and intervention. Participants used a wearable device for PTAS application at home, the Mobile Health Systems Lab (MHSL) SleepBand v3 (MHSL-SB)^40,41^. Participants were switched from three consecutive nights of PTAS (STIM) to three nights of sham stimulation (SHAM), or vice versa, after a washout period of four nights (Figure 1 a). Thus, both intervention periods took place on the same weekdays. During interventions, participants wore the MHSL-SB the entire night at their regular bedtimes in their regular sleeping environment. The experimenter instructed all participants to maintain a regular sleep– wake rhythm and maintain their habitual routines before and throughout the entire study. Adherence was assessed by regular telephone calls with the experimenter scheduled at least before, after, and once during each intervention period; daily questionnaires; and wrist actigraphy worn by the participants seven days before and throughout the entire study.

With two participants, we had to repeat the STIM intervention because more than one night was not recorded correctly due to technical failures in the initial intervention period. We had to exclude two participants after completing both interventions due to restless legs syndrome and sleep apnea. One participant was excluded from each subgroup. In the final analysis, we included 21 patients, with 117 intervention nights available out of 126. Nine nights were unavailable for analyses owing to technical failures: six in SHAM and three in STIM.

### Participants

Participants were recruited via advertisements in institutional websites, printed magazines of the Parkinson Schweiz organization, patient support groups, the movement disorder outpatient clinic of the Department of Neurology, University Hospital Zurich, and selected neurological practices in Switzerland.

Screening included a visit followed by up to four screening nights with the MHSL-SB. The consultation took place at the study site or online and at the participants’ homes and included detailed history taking and medical and neurological examination by a study physician with experience in PD and sleep medicine (SJS or JH). Motor examination was performed in the medical ON state after intake of regular dopaminergic medication for documentation of the motor subtype and Movement Disorders Society Unified Parkinson’s Disease Rating Scale Part III (UPDRS III)^61^. Participants completed standardized questionnaires, including PDSS-2, Epworth Sleepiness Scale (ESS), RBD-Single-Question Screen (RBD-SQ)^62^, and the Hamilton Depression Scale (HADS) and provided information about symptoms of restless legs syndrome and risk factors of obstructive sleep apnea.

Eligible participants had to have a diagnosis of PD according to international criteria^63^, with mild to moderate disease severity (Hoehn & Yahr Stages II-III), and self-reported sleep disturbance defined by ≥7 points on questions 1–3 and 14 of the Parkinson’s Disease Sleep Scale revised version (PDSS-2)^50,64^. The PDSS-2 is a self-report questionnaire composed of three factors covering [1] motor symptoms at night, [2] PD symptoms at night, and [3] disturbed sleep, from which we summed items 1–3 and 14 (bad sleep quality, difficulties falling asleep, difficulties staying asleep, tired and sleepy after waking in the morning)^50^. We chose this approach to select participants with the most common phenotype of disturbed sleep in PD^65,66^. Likewise, we excluded patients with sleep disorders that can significantly disrupt sleep and are common outside PD, such as sleep apnea with apnea-hypopnea index (AHI) > 15/h and restless legs syndrome, but not with sleep disturbances more closely related to PD, such as REM sleep behavior disorder and sleep disturbances rated with the PDSS-2^65,66^. The most important further additional inclusion criteria were the ability to follow study procedures and stable dosing of Parkinsonian medication for at least 14 days prior to the first intervention and during the entire study. The most important exclusion criteria were known presence of neurologic (other than PD), psychiatric, or severe medical conditions, atypical or poorly levodopa-responsive Parkinsonism, cognitive impairment (Montréal Cognitive Assessment, MoCA<24), regular use of central nervous system depressant or sleep-inducing substances, working night shifts, travelling more than two time zones in the last month before or during intervention, alcohol abuse (i.e. >0.5 l wine or 1 l beer per day), substance abuse, high caffeine consumption (>5 servings per day, including coffee and energy drinks), implants for deep brain stimulation, inability to hear the tones of the MHSL-SB, and absence of PTAS-induced SWA enhancement. The total numbers number of screened and included subjects can be seen in Figure 2. Although subjectively disturbed sleep, with symptoms of insomnia, was an inclusion criterion, we had to exclude two participants due to worsening of sleep-onset insomnia when testing the device at home. Two participants, one from each subgroup, had to be excluded from the analysis after completing the study, because all criteria for restless legs syndrome were met (*n* = 1), and positional sleep apnea was suspected and later confirmed (*n* = 1) only towards the end of the intervention.

Screening nights took place at the participants’ homes. The experimenters (SJS, JH, LK) provided participants with the MHSL-SB, a tablet, and instructions on usage. From the data collected during screening nights, we determined eligibility for applying the device at home and the occurrence of PTAS-induced increase in SWA and titrated the volume of the MHSL-SB if needed.

### Phase-targeted auditory stimulation

Phase-targeted auditory stimulation was applied with the MHSL-SB as previously described in detail^41^. In brief, the MHSL-SB is a configurable wearable system for sleep recording and modulation that enables real-time EEG processing and enhancement of slow waves with PTAS^35,40,41^, which can be optimized for use in PD patients^59^. The system records single channel EEG at the forehead (Fpz) referenced to the mastoid for real-time signal processing, eye movements (two electrooculography, EOG, channels), and muscle activity (two electromyography, EMG, channels) at the chin for post-processing purposes.

As in previous studies, we used alternating 6-s windows during which stimulation is, in principle, enabled (ON) or disabled (OFF) ^41^. The system delivers nonarousing 50 ms stimuli of multifrequency pink noise targeting the up-phase of slow waves using a phase-locked loop (PLL; target phase of 45 degrees) if spectral power threshold criteria for NREM sleep and slow wave detection are met and NREM is considered stable (continuous NREM sleep for 10 minutes for the first NREM sleep event and for 3 minutes for any following events). To achieve sufficiently strong stimuli while avoiding arousal induced by the stimuli, the system automatically adapts the volume during PTAS within a predefined dB range: the volume increases within predefined limits during deep sleep in the absence of arousals or decreases or pauses in the presence of arousals detected by the system. In this study, we set 52 dB as default, tested this volume during wakefulness, and titrated up to a range of 70–80 dB if needed to surpass individual hearing thresholds. In addition, we lowered the preset volume if participants reported hearing tones during the night. Applied volumes ranged between minimal 39.4 to 57.7 (median 42.8) dB and maximal 57.6 to 68.6 (median 64.9) dB.

During SHAM, all procedures were identical to the STIM condition, but the device volume was muted while recording the signal along with timing and number of theoretical stimuli that would have been applied under STIM conditions.

Previous studies applying PTAS without individualizing volumes and algorithm thresholds for spectral power or amplitude showed that the degree of SWA enhancement varies greatly and can be absent in old-aged adults^44,45,59^. Because our aim was to investigate the potential of PTAS when SWA enhancement is present, we individualized settings for volume and amplitude thresholds during screening nights to achieve a quantifiable PTAS-induced increase in SWA prior to the intervention. Five out of 33 participants (15 %) had to be excluded owing to a lack of response despite the adaptation of settings (Figure 2).

Finally, we implemented two protocols with different durations of PTAS: <u>Whole-night PTAS:</u> PTAS was applied for the entire night, as performed in the study using MHSL-SB in healthy older adults ^41^. <u>Part-night PTAS:</u> While wearing the device for the entire night PTAS was restricted to the deep sleep-rich, first part of the night: 2.5 hours after the first stimulus in all but one participant who had low numbers of stimuli due to sleep fragmentation and thus received 3 hours after the first stimulus. We switched between these protocols halfway after completing the first 14 participants with whole-night PTAS (posthoc defined as the whole-night subgroup) to complete the following nine participants with part-night PTAS (posthoc defined as the part-night subgroup). This switch was prompted by new findings that whole-night PTAS may affect mood through its impact on REM sleep^41^. Accordingly, the part-night PTAS intends to align with the physiological predominance of deep sleep during the first part of the night and avoids potential impact on REM sleep, which physiologically predominates during the second part of the night. In the whole-night PTAS protocol, on average (±SD), 2037.92 ± 776.41 stimuli were applied per night; in the part-night protocol, 1589.96 ± 967.71 stimuli.

### Sleep macrostructure

The MHSL-SB single-channel EEG (FpZ-A2), EOG, and chin EMG were used for scoring sleep stages for all intervention nights. Experienced sleep specialists (SF, PV, RP, SJS) scored according to the AASM criteria^67^. In the whole-night subgroup, initial automated scoring used a deep learning classification^41^ that had been retrained on a PD data set, followed by expert control, which took precedent in case of disagreement with the algorithm. In the part-night subgroup, only expert scoring was performed. From scored data, we calculated absolute and relative amounts of sleep stages (N1, N2, N3, R), duration and incidences of wake after sleep onset (WASO), number of awakenings, and sleep efficiency (SE, % TST of sleep period time).

### EEG analysis

EEG was analyzed as previously described^41^. In brief, EEG signals were high- and low-pass filtered at 0.5 Hz and 40 Hz, respectively, with the EEGLAB toolbox for Matlab (The MathWorks, Inc., Natick, MA)^68,69^. Then we used the pwelch function to calculate EEG spectral power between 0.5 and 30 Hz for all 6-s ON and OFF windows. We only included windows that were classified as NREM sleep without artifacts by the online algorithm and were offline scored as N2 or N3. Furthermore, we applied a semi-automated artifact rejection and automated outlier detection using the *isoutlier* function of Matlab^70,71^.

To be consistent with Lustenberger et al. (2022), we normalized the spectral power in each 0.25 Hz frequency bin to the summed power up to 30 Hz for the spectral density analysis (Figure 3a, 3b) ^41^. In line with their approach, we calculated the absolute power in the SWA range in ON and OFF windows that were grouped by the number of stimuli (#stimuli) during ON and the corresponding OFF windows (1–2, 3–4, >4) to assess a potential dose-dependent response to PTAS (Figure 3c). However, because spectral density analysis had revealed effects in a broader SWA range, we did not restrict this analysis to the low low-frequency range of SWA but included the full SWA band: 0.75–4.5 Hz. Furthermore, we also categorized OFF windows by the number of stimuli during preceding ON windows to assess whether preceding stimulation influenced subsequent OFF windows in a dose-dependent manner.

### Subjective outcomes

Participants answered visual analogue scales (VAS) for several symptoms and the PDSS-2 on a tablet. The PDSS-2 rates the severity of 15 typical nocturnal symptoms in PD by asking about their frequencies over one week (never, 1d/week, 2-3d/week, 4-5d/week, >5d/week)^50^. We adapted the scale to assess symptoms over three nights, the duration of our intervention, instead of one week (0/3 nights, 1/3 nights, 2/3 nights, 3/3 nights). This adapted PDSS-2 was collected before and after each intervention (Pre, Post), to assess change during intervention (Figure 1 a). We considered the total PDSSS-2 score and items 1-3 and 14 from the disturbed sleep subscale^50^.

Daily subjective ratings included VAS (0–100) for subjective sleep quality (how refreshing was your sleep? How good was your sleep quality? every morning), mood (every evening), and average daytime sleepiness (every evening), and the Karolinska Sleepiness Scale for assessing momentary sleepiness (every morning and evening). These outcomes were collected every day during interventions, and on that day on which the intervention started as a baseline (pre).

### Statistical analysis

The statistical analysis was conducted using R Studio software (R Foundation for Statistical Computing, Vienna, Austria). We ran a robust linear mixed model (RLMM) applying the Kenword-Roger approximation method^72^ to compare the effects of STIM to SHAM on sleep (NREM sleep EEG power, sleep macrostructure) and subjective outcomes. In these models, we included condition (STIM vs. SHAM), period (1 vs. 2), sequence (STIM–SHAM vs. SHAM– STIM), and in case of combined analyses, subgroup (whole-night vs. part-night) as fixed factors and subject and condition as nested random factor. For the dose–response analysis of SWA in ON and OFF windows (Fig 3c), we averaged values across the nights and included #stimuli (1– 2, 3–4, >4) and its interaction with the condition factor as fixed factors.

For the PDSS-2 analysis, we included the time point factor (pre- vs. postintervention) and its interaction with the condition factor to assess the change induced by the intervention. For all other analyses, we included the night (1, 2, 3) fixed factor in the models. Because previous studies suggested that the beneficial effects of PTAS might require several nights of intervention^38,39^, we also tested for interaction effects between condition and night in subjective outcomes. Following any significant interaction, we ran separate analyses for categories with different numbers of stimuli, time points, or nights. In the daily assessed subjective ratings, we further included the corresponding baseline value (pre) as a predictor. Any *p*-values <0.05 were considered significant.

## Acknowledgements

We are especially grateful to all participants of this study. We further thank the funding agencies Swiss National Science Foundations (grant numbers 188790 to CRB, and PZ00P3_179795 to CL), Parkinson Schweiz, Schweizerische Hirnstiftung, and Swiss Neurological Society (stipend to SJS) for their generous support. We thank Neil Rupprecht, Mario Andina, Silvia Haug, Nadja Frei, Alessandra Pfister, and Valentino Mutschler, for supporting this study as part of their studies at the University of Zurich and ETH Zurich, Switzerland. We thank Stephanie Huwyler for technical assistance, Esther Werth for support of the manual sleep analysis, and the entire SleepLoop Consortium. This study was conducted as part of the Hochschulmedizin Zurich Flagship Project SleepLoop (www.sleeploop.ch).

## Author contributions

SJS, RH, CRB, and AM conceptualized and designed the study. SJS, JH, and LK prepared and conducted the study under supervision of AM and CRB. RH, WK, GD, LF, CL, RS, VJ, and MS provided methodological and/or technical support and guidance. LB provided the retrained automatic scoring algorithm. SJS, SF, RP, and PV visually scored the sleep EEG data. SJS, JH, SF, KR, and AM analyzed data. SJS wrote and AM revised the main manuscript text, all authors reviewed the manuscript and provided feedback and approved the submitted version of the manuscript.

## Data availability

Data can be requested from the corresponding author upon reasonable request and in accordance with regulatory rules.

## References

1 Pringsheim, T., Jette, N., Frolkis, A. & Steeves, T. D. The prevalence of Parkinson’s disease: A systematic review and meta-analysis. Movement disorders 29, 1583–1590 (2014).

2 Dorsey, E. R. & Bloem, B. R. The Parkinson Pandemic—A Call to ActionThe Parkinson PandemicThe Parkinson Pandemic. JAMA neurology 75, 9–10, doi:10.1001/jamaneurol.2017.3299 (2018).

3 Feigin, V. L. et al. Global, regional, and national burden of neurological disorders during 1990–2015: a systematic analysis for the Global Burden of Disease Study 2015. The Lancet Neurology 16, 877–897 (2017).

4 Martinez-Martin, P., Rodriguez-Blazquez, C., Kurtis, M. M., Chaudhuri, K. R. & on Behalf of the, N. V. G. The impact of non-motor symptoms on health-related quality of life of patients with Parkinson’s disease. Movement Disorders 26, 399–406, doi:10.1002/mds.23462 (2011).

5 Stefani, A. & Högl, B. Sleep in Parkinson’s disease. Neuropsychopharmacology : official publication of the American College of Neuropsychopharmacology 45, 121–128, doi:10.1038/s41386-019-0448-y (2020).

6 Ju, Y. E., Lucey, B. P. & Holtzman, D. M. Sleep and Alzheimer disease pathology--a bidirectional relationship. Nature reviews. Neurology 10, 115–119, doi:10.1038/nrneurol.2013.269 (2014).

7 Mander, B. A., Winer, J. R., Jagust, W. J. & Walker, M. P. Sleep: A Novel Mechanistic Pathway, Biomarker, and Treatment Target in the Pathology of Alzheimer’s Disease? Trends in neurosciences 39, 552–566, doi:10.1016/j.tins.2016.05.002 (2016).

8 Bellesi, M., Riedner, B. A., Garcia-Molina, G. N., Cirelli, C. & Tononi, G. Enhancement of sleep slow waves: underlying mechanisms and practical consequences. Frontiers in systems neuroscience 8, doi:10.3389/fnsys.2014.00208 (2014).

9 Malkani, R. G. & Zee, P. C. Brain Stimulation for Improving Sleep and Memory. Sleep medicine clinics 15, 101–115, doi:10.1016/j.jsmc.2019.11.002 (2020).

10 Lee, Y. F., Gerashchenko, D., Timofeev, I., Bacskai, B. J. & Kastanenka, K. V. Slow Wave Sleep Is a Promising Intervention Target for Alzheimer’s Disease. Front Neurosci 14, 705, doi:10.3389/fnins.2020.00705 (2020).

11 Vyazovskiy, V. V. & Harris, K. D. Sleep and the single neuron: the role of global slow oscillations in individual cell rest. Nature Reviews Neuroscience 14, 443–451, doi:10.1038/nrn3494 (2013).

12 Gorgoni, M. & Galbiati, A. Non-REM sleep electrophysiology in REM sleep behaviour disorder: A narrative mini-review. Neuroscience & Biobehavioral Reviews 142, 104909, 10.1016/j.neubiorev.2022.104909 (2022).

13 Sunwoo, J.-S. et al. NREM Sleep EEG Oscillations in Idiopathic REM Sleep Behavior Disorder: A study of sleep spindles and slow oscillations. Sleep, doi:10.1093/sleep/zsaa160 (2020).

14 Brunner, H. et al. Microstructure of the non-rapid eye movement sleep electroencephalogram in patients with newly diagnosed Parkinson’s disease: effects of dopaminergic treatment. Mov Disord 17, 928–933, doi:10.1002/mds.10242 (2002).

15 Diederich, N. J., Vaillant, M., Mancuso, G., Lyen, P. & Tiete, J. Progressive sleep ’destructuring’ in Parkinson’s disease. A polysomnographic study in 46 patients. *Sleep Med* **6**, 313-318, doi:10.1016/j.sleep.2005.03.011 (2005).

16 Davin, A. et al. Early onset of sleep/wake disturbances in a progressive macaque model of Parkinson’s disease. Sci Rep 12, 17499, doi:10.1038/s41598-022-22381-z (2022).

17 Schreiner, S. J. et al. Sleep spindle and slow wave activity in Parkinson disease with excessive daytime sleepiness. Sleep, doi:10.1093/sleep/zsac165 (2022).

18 Amato, N. et al. Levodopa-induced dyskinesia in Parkinson disease: Sleep matters. Ann Neurol 84, 905–917, doi:10.1002/ana.25360 (2018).

19 Wood, K. H. et al. Slow Wave Sleep and EEG Delta Spectral Power are Associated with Cognitive Function in Parkinson’s Disease. Journal of Parkinson’s disease, doi:10.3233/jpd-202215 (2020).

20 Schreiner, S. J. et al. Reduced Regional NREM Sleep Slow-Wave Activity Is Associated With Cognitive Impairment in Parkinson Disease. Frontiers in neurology 12, doi:10.3389/fneur.2021.618101 (2021).

21 Meinhold, L. et al. T2 MRI visible Perivascular Spaces in Parkinson’s Disease: clinical significance and association with polysomnography measured sleep. Sleep, doi:10.1093/sleep/zsae233 (2024).

22 Schreiner, S. J. et al. Slow-wave sleep and motor progression in Parkinson disease. Ann Neurol 85, 765–770, doi:10.1002/ana.25459 (2019).

23 Bugalho, P. et al. Polysomnographic predictors of sleep, motor and cognitive dysfunction progression in Parkinson’s Disease: a longitudinal study. Sleep Medicine, 10.1016/j.sleep.2020.06.020 (2020).

24 Chen, J. et al. Correlation of slow-wave sleep with motor and nonmotor progression in Parkinson’s disease. Annals of Clinical and Translational Neurology 11, 554–563, 10.1002/acn3.51975 (2024).

25 Tao, M.-X. et al. Slow-wave sleep and REM sleep without atonia predict motor progression in Parkinson’s disease. Sleep Medicine 115, 155–161, 10.1016/j.sleep.2024.02.003 (2024).

26 Büchele, F., Hackius, M., Schreglmann, S. R. &, et al. Sodium oxybate for excessive daytime sleepiness and sleep disturbance in parkinson disease: A randomized clinical trial. JAMA neurology 75, 114–118, doi:10.1001/jamaneurol.2017.3171 (2018).

27 Morawska, M. M. et al. Slow-wave sleep affects synucleinopathy and regulates proteostatic processes in mouse models of Parkinson’s disease. Sci Transl Med 13, eabe7099, doi:10.1126/scitranslmed.abe7099 (2021).

28 Xie, L. et al. Sleep drives metabolite clearance from the adult brain. Science 342, 373–377, doi:10.1126/science.1241224 (2013).

29 Holth, J. K. et al. The sleep-wake cycle regulates brain interstitial fluid tau in mice and CSF tau in humans. *Science*, eaav2546, doi:10.1126/science.aav2546 (2019).

30 Kang, J. E. et al. Amyloid-beta dynamics are regulated by orexin and the sleep-wake cycle. Science 326, 1005–1007, doi:10.1126/science.1180962 (2009).

31 Ju, Y.-E. S. et al. Slow wave sleep disruption increases cerebrospinal fluid amyloid-β levels. Brain 140, 2104–2111, doi:10.1093/brain/awx148 (2017).

32 Olsson, M., Arlig, J., Hedner, J., Blennow, K. & Zetterberg, H. Sleep deprivation and cerebrospinal fluid biomarkers for Alzheimer’s disease. Sleep 41, doi:10.1093/sleep/zsy025 (2018).

33 Feher, K. D. et al. Shaping the slow waves of sleep: A systematic and integrative review of sleep slow wave modulation in humans using non-invasive brain stimulation. Sleep medicine reviews 58, 101438, doi:10.1016/j.smrv.2021.101438 (2021).

34 Ngo, H.-Viet V., Martinetz, T., Born, J. & Mölle, M. Auditory Closed-Loop Stimulation of the Sleep Slow Oscillation Enhances Memory. Neuron 78, 545–553, 10.1016/j.neuron.2013.03.006 (2013).

35 Fattinger, S. et al. Deep sleep maintains learning efficiency of the human brain. Nature Communications 8, 15405, doi:10.1038/ncomms15405 (2017).

36 Lustenberger, C. et al. High-density EEG characterization of brain responses to auditory rhythmic stimuli during wakefulness and NREM sleep. NeuroImage 169, 57–68, 10.1016/j.neuroimage.2017.12.007 (2018).

37 Besedovsky, L. et al. Auditory closed-loop stimulation of EEG slow oscillations strengthens sleep and signs of its immune-supportive function. Nature Communications 8, 1984, doi:10.1038/s41467-017-02170-3 (2017).

38 Wunderlin, M. et al. Acoustic stimulation during sleep predicts long-lasting increases in memory performance and beneficial amyloid response in older adults. Age and Ageing 52, doi:10.1093/ageing/afad228 (2023).

39 Zeller, C. J. et al. Multi-night acoustic stimulation is associated with better sleep, amyloid dynamics, and memory in older adults with cognitive impairment. GeroScience, doi:10.1007/s11357-024-01195-z (2024).

40 Ferster, M. L., Lustenberger, C. & Karlen, W. Configurable Mobile System for Autonomous High-Quality Sleep Monitoring and Closed-Loop Acoustic Stimulation. IEEE Sensors Letters 3, 1–4, doi:10.1109/LSENS.2019.2914425 (2019).

41 Lustenberger, C. et al. Auditory deep sleep stimulation in older adults at home: a randomized crossover trial. Commun Medicine 2, 30, doi:10.1038/s43856-022-00096-6 (2022).

42 Zeller, C. J., Züst, M. A., Wunderlin, M., Nissen, C. & Klöppel, S. The promise of portable remote auditory stimulation tools to enhance slow-wave sleep and prevent cognitive decline. J Sleep Res, e13818, doi:10.1111/jsr.13818 (2023).

43 Papalambros, N. A. et al. Acoustic enhancement of sleep slow oscillations in mild cognitive impairment. Annals of clinical and translational neurology 6, 1191–1201, doi:10.1002/acn3.796 (2019).

44 Papalambros, N. A. et al. Acoustic Enhancement of Sleep Slow Oscillations and Concomitant Memory Improvement in Older Adults. Front Hum Neurosci 11, 109, doi:10.3389/fnhum.2017.00109 (2017).

45 Schneider, J., Lewis, P. A., Koester, D., Born, J. & Ngo, H. V. Susceptibility to auditory closed-loop stimulation of sleep slow oscillations changes with age. Sleep, doi:10.1093/sleep/zsaa111 (2020).

46 Mizrahi-Kliger, A. D., Kaplan, A., Israel, Z., Deffains, M. & Bergman, H. Basal ganglia beta oscillations during sleep underlie Parkinsonian insomnia. Proceedings of the National Academy of Sciences 117, 17359–17368, doi:doi:10.1073/pnas.2001560117 (2020).

47 Zahed, H. et al. The Neurophysiology of Sleep in Parkinson’s Disease. Movement Disorders 36, 1526–1542, 10.1002/mds.28562 (2021).

48 Zhang, Y. et al. Sleep in Parkinson’s disease: A systematic review and meta-analysis of polysomnographic findings. Sleep medicine reviews 51, 101281, doi:10.1016/j.smrv.2020.101281 (2020).

49 Memon, A. A. et al. Quantitative Sleep Electroencephalogram in Parkinson’s Disease: A Case-Control Study. Journal of Parkinson’s disease 13, 351–365, doi:10.3233/jpd-223565 (2023).

50 Trenkwalder, C. et al. Parkinson’s disease sleep scale—validation of the revised version PDSS-2. Movement Disorders 26, 644–652, doi:doi:10.1002/mds.23476 (2011).

51 Winer, J. R. et al. Sleep Disturbance Forecasts &#x3b2;-Amyloid Accumulation across Subsequent Years. Current Biology, doi:10.1016/j.cub.2020.08.017 (2020).

52 Ngo, H. V., Claassen, J. & Dresler, M. Sleep: Slow Wave Activity Predicts Amyloid-β Accumulation. Current biology : CB 30, R1371–r1373, doi:10.1016/j.cub.2020.09.058 (2020).

53 Stanyer, E. C. et al. The impact of acoustic stimulation during sleep on memory and sleep architecture: A meta-analysis. J Sleep Res 31, e13385, doi:10.1111/jsr.13385 (2022).

54 Andre, C. et al. Rapid Eye Movement Sleep, Neurodegeneration, and Amyloid Deposition in Aging. Annals of neurology 93, 979–990, doi:10.1002/ana.26604 (2023).

55 Arnulf, I. et al. Parkinson’s disease and sleepiness: an integral part of PD. Neurology 58, 1019–1024 (2002).

56 Bogan, R. K., Roth, T., Schwartz, J. & Miloslavsky, M. Time to response with sodium oxybate for the treatment of excessive daytime sleepiness and cataplexy in patients with narcolepsy. Journal of clinical sleep medicine : JCSM : official publication of the American Academy of Sleep Medicine 11, 427–432, doi:10.5664/jcsm.4598 (2015).

57 Van den Bulcke, L., et al. Acoustic Stimulation to Improve Slow-Wave Sleep in Alzheimer’s Disease: A Multiple Night At-Home Intervention. The American Journal of Geriatric Psychiatry, 10.1016/j.jagp.2024.07.002 (2024).

58 Wunderlin, M., Koenig, T., Zeller, C., Nissen, C. & Züst, M. A. Automatized online prediction of slow-wave peaks during non-rapid eye movement sleep in young and old individuals: Why we should not always rely on amplitude thresholds. Journal of Sleep Research 31, e13584, 10.1111/jsr.13584 (2022).

59 Ferster, M. L. et al. Benchmarking real-time algorithms for in-phase auditory stimulation of low amplitude slow waves with wearable EEG devices during sleep. IEEE Transactions on Biomedical Engineering, 1–1, doi:10.1109/TBME.2022.3157468 (2022).

60 Krugliakova, E. et al. Exploring the local field potential signal from the subthalamic nucleus for phase-targeted auditory stimulation in Parkinson’s disease. Brain Stimul 17, 769–779, doi:10.1016/j.brs.2024.06.007 (2024).

61 Goetz, C. G. et al. Movement Disorder Society-sponsored revision of the Unified Parkinson’s Disease Rating Scale (MDS-UPDRS): Process, format, and clinimetric testing plan. Movement Disorders 22, 41–47, 10.1002/mds.21198 (2007).

62 Postuma, R. B. et al. A single-question screen for rapid eye movement sleep behavior disorder: a multicenter validation study. Mov Disord 27, 913–916, doi:10.1002/mds.25037 (2012).

63 Postuma, R. B. et al. MDS clinical diagnostic criteria for Parkinson’s disease. Movement Disorders 30, 1591–1601, doi:10.1002/mds.26424 (2015).

64 Suzuki, K. et al. Evaluation of cutoff scores for the Parkinson’s disease sleep scale-2. Acta neurologica Scandinavica 131, 426–430, doi:10.1111/ane.12347 (2015).

65 Chahine, L. M., Amara, A. W. & Videnovic, A. A systematic review of the literature on disorders of sleep and wakefulness in Parkinson’s disease from 2005 to 2015. Sleep medicine reviews 35, 33–50, doi:10.1016/j.smrv.2016.08.001 (2017).

66 Iranzo, A., Cochen De Cock, V., Fantini, M. L., Pérez-Carbonell, L. & Trotti, L. M. Sleep and sleep disorders in people with Parkinson’s disease. The Lancet Neurology 23, 925–937, doi:10.1016/S1474-4422(24)00170-4 (2024).

67 Medicine, A. A. o. S. *International classification of sleep disorders*. (Darien, IL, 2014).

68 Brunner, C., Delorme, A. & Makeig, S. Eeglab - an Open Source Matlab Toolbox for Electrophysiological Research. Biomed Tech (Berl) 58 **Suppl 1**, doi:10.1515/bmt-2013-4182 (2013).

69 Delorme, A. & Makeig, S. EEGLAB: an open source toolbox for analysis of single-trial EEG dynamics including independent component analysis. J Neurosci Methods 134, 9–21, doi:10.1016/j.jneumeth.2003.10.009 (2004).

70 Huber, R. et al. Exposure to pulsed high-frequency electromagnetic field during waking affects human sleep EEG. Neuroreport 11, 3321–3325, doi:10.1097/00001756-200010200-00012 (2000).

71 Lustenberger, C., Wehrle, F., Tüshaus, L., Achermann, P. & Huber, R. The Multidimensional Aspects of Sleep Spindles and Their Relationship to Word-Pair Memory Consolidation. Sleep 38, 1093–1103, doi:10.5665/sleep.4820 (2015).

72 Halekoh, U. & Højsgaard, S. A Kenward-Roger Approximation and Parametric Bootstrap Methods for Tests in Linear Mixed Models - The R Package pbkrtest. Journal of Statistical Software 59, doi:10.18637/jss.v059.i09 (2014).

