## Supplementary tables for "Auditory Enhancement of Sleep Slow Waves in People with Parkinson’s Disease: A Proof-of-Concept Study"

**Table S1: Patient characteristics across subgroups**

| | Whole-night PTAS, n = 13 | Part-night PTAS, n = 8 | p-value |
|---|---|---|---|
| Age (years) | 54 (50, 59) | 59 (56, 62) | 0.20 |
| Females | 9 (69%) | 1 (13%) | 0.02 |
| BMI (kg/m2) | 26.05 (24.86, 27.64) | 25.52 (23.69, 26.62) | 0.50 |
| Disease duration (years) | 6 (4, 8) | 5 (3.75, 6) | 0.29 |
| Motor Subtype: | | | 0.80 |
| Akinetic rigid | 8 (62%) | 6 (75%) | |
| Mixed | 3 (23%) | 2 (25%) | |
| Tremor-dominant | 2 (15%) | 0 (0%) | |
| Hoehn & Yahr Stage: | | | 0.40 |
| II | 13 (100%) | 7 (88%) | |
| III | 0 (0%) | 1 (13%) | |
| Available DAT scan | 7 (54%) | 3 (38%) | 0.69 |
| Levodopa equivalent dose (mg/d) | 595 (325, 780) | 772 (616, 884) | 0.20 |
| MDS-UPDRS III | 19 (16, 23) | 28 (22, 34) | 0.05 |
| Montréal Cognitive Assessment | 27 (27, 29) | 29 (26.75, 29.25) | 0.50 |
| PDSS-2 | 17 (15, 21) | 19.5 (15.0, 25.3) | 0.60 |
| PDSS-2 disturbed sleep subscale | 10 (9, 11) | 8.00 (7.00, 9.75) | 0.20 |
| Epworth Sleepiness Scale | 12 (9, 14) | 9.00 (7.75, 12.50) | 0.30 |
| Sleep quality (VAS) | 34 (31, 36) | 62 (38, 63) | 0.02 |
| STOP-BANG OSA | | | 0.30 |
| 1 | 8 (62%) | 3 (38%) | |
| 2 | 3 (23%) | 5 (63%) | |
| 3 | 2 (15%) | 0 (0%) | |
| RBD Single-Question Screen | | | >0.9 |
| No | 9 (69%) | 5 (63%) | |
| Yes | 4 (31%) | 3 (38%) | |

Median (IQR); Wilcoxon ranksum or Chi2 test.

DAT = dopamine transporter; BMI, Body Mass Index; PDSS-2, Parkinson's Diseases Sleep Scale-2

RBD, Rapid eye movement sleep behavior disorder; MDS-UPDRS, Movement Disorder Society Unified Parkinson's Disease Rating Scale; VAS, Visual analogue scale; OSA, Obstructive sleep apnea

**Table S2: Sleep macrostructure during first part of the night**

| *n* = 21 | STIM | | SHAM | | P condition |
|---|---|---|---|---|---|
| | Mean | SD | Mean | SD | |
| WASO (min) | 11.09 | 10.82 | 13.47 | 13.64 | 0.46 |
| Awakenings (n) | 10.48 | 8.91 | 10.33 | 6.98 | 0.50 |
| SE (%) | 95.00 | 6.16 | 94.59 | 6.85 | 0.85 |
| TST (min) | 166.23 | 13.39 | 167.68 | 11.05 | 0.45 |
| N1 (min) | 11.77 | 5.18 | 11.29 | 3.38 | 0.75 |
| N2 (min) | 85.78 | 27.02 | 91.01 | 22.23 | 0.50 |
| N3 (min) | 51.36 | 30.29 | 51.30 | 16.74 | 0.66 |
| R (min) | 17.32 | 8.25 | 14.07 | 8.85 | 0.49 |
| N1 (%) | 7.16 | 3.30 | 6.72 | 1.77 | 0.61 |
| N2 (%) | 51.33 | 15.49 | 53.76 | 11.34 | 0.47 |
| N3 (%) | 30.87 | 17.19 | 31.03 | 10.78 | 0.64 |
| R (%) | 10.64 | 5.13 | 8.49 | 5.29 | 0.37 |

P shows p-values from robust linear mixed models (see methods section).

WASO, wake after sleep onset; SE, sleep efficiency; TST, total sleep time; N1-3, non-rapid eye movement sleep stage 1-3; R, rapid eye movement sleep; % are expressed relative to TST

**Table S3: Sleep macrostructure during whole-night**

| **Whole-night PTAS subgroup** *n* = 13 | **STIM** | | **SHAM** | | **P condition** |
|---|---|---|---|---|---|
| | Mean | SD | Mean | SD | |
| WASO (min) | 64.58 | 50.02 | 49.04 | 33.13 | 0.06 |
| Awakenings (n) | 42.45 | 33.06 | 39.87 | 28.11 | 0.42 |
| SE (%) | 84.84 | 10.97 | 88.53 | 8.29 | 0.05 |
| TST (min) | 347.29 | 63.37 | 368.65 | 51.25 | 0.12 |
| N1 (min) | 34.85 | 10.73 | 32.65 | 9.66 | 0.23 |
| N2 (min) | 193.83 | 42.53 | 204.38 | 33.52 | 0.25 |

| | | | | | |
|---|---|---|---|---|---|
| N3 (min) | 50.92 | 19.64 | 59.41 | 24.69 | 0.21 |
| R (min) | 67.70 | 36.88 | 72.21 | 35.49 | 0.44 |
| N1 (%) | 22.41 | 11.65 | 21.86 | 8.81 | 0.87 |
| N2 (%) | 55.73 | 7.03 | 55.37 | 7.40 | 0.99 |
| N3 (%) | 15.19 | 6.14 | 16.29 | 6.05 | 0.53 |
| R (%) | 18.75 | 8.20 | 19.30 | 8.08 | 0.63 |
| **Part-night PTAS subgroup** *n* = 8 | **STIM** | | **SHAM** | | **P condition** |
| | **Mean** | **SD** | **Mean** | **SD** | |
| WASO (min) | 51.42 | 14.99 | 57.81 | 25.81 | 0.48 |
| Awakenings (n) | 36.00 | 10.31 | 39.46 | 7.25 | 0.49 |
| SE (%) | 87.43 | 3.86 | 86.57 | 4.56 | 0.36 |
| TST (min) | 347.15 | 50.02 | 352.53 | 50.46 | 0.59 |
| N1 (min) | 30.11 | 12.26 | 32.46 | 13.64 | 0.70 |
| N2 (min) | 173.93 | 59.99 | 183.89 | 45.62 | 0.46 |
| N3 (min) | 85.46 | 45.71 | 83.71 | 26.46 | 0.52 |
| R (min) | 57.65 | 13.08 | 52.47 | 18.77 | 0.49 |
| N1 (%) | 18.33 | 11.44 | 17.83 | 11.88 | 0.60 |
| N2 (%) | 49.21 | 13.60 | 51.66 | 9.02 | 0.25 |
| N3 (%) | 25.55 | 13.50 | 24.39 | 7.58 | 0.85 |
| R (%) | 16.62 | 3.71 | 14.80 | 5.22 | 0.32 |

P shows p-values from robust linear mixed models (see methods section).

WASO, wake after sleep onset; SE, sleep efficiency; TST, total sleep time; N1-3, non-rapid eye movement sleep stage 1-3; R, rapid eye movement sleep; % are expressed relative to TST

**Table S4: Subjective outcomes**

| Whole-night subgroup, *n* = 13 | STIM | | | SHAM | | | | |
|---|---|---|---|---|---|---|---|---|
| | Median | Min | Max | Median | Min | Max | P condition | P condition x night |
| VAS Sleep Quality | 3 | -62 | 47 | 1 | -63 | 50 | 0.287 | 0.15 |

| | | | | | | | | |
|---|---|---|---|---|---|---|---|---|
| VAS Sleep Refreshment | 4 | -33 | 45 | 11 | -19 | 44 | 0.405 | 0.22 |
| KSS Morning | 0 | -7 | 5 | 0 | -7 | 4 | 0.1 | 0.13 |
| KSS Evening | 0 | -4 | 3 | -1 | -6 | 2 | 0.091 | 1.00 |
| VAS Mood | 5.5 | -33 | 44 | 0 | -24 | 40 | 0.155 | 0.36 |
| VAS Daytime Sleepiness | -4 | -34 | 33 | -6.5 | -34 | 23 | 0.246 | 0.31 |
| PDSS-2 Disturbed Sleep | -1 | -4 | 5 | 1 | -4 | 6 | 0.89* | NA |
| PDSS-2 Total | 0 | -4 | 5 | 1 | -4 | 6 | 0.902* | NA |
| Part-night subgroup, *n* = 8 | STIM | | | SHAM | | | | |
| | Median | Min | Max | Median | Min | Max | P condition | P condition x night |
| VAS Sleep Quality | -5 | -61 | 19 | -8 | -75 | 28 | 0.265 | 0.46 |
| VAS Sleep Refreshment | -0.5 | -40 | 20 | -9 | -49 | 33 | 0.203 | 0.42 |
| KSS Morning | 0 | -2 | 2 | 0 | -3 | 2 | 0.285 | 0.92 |
| KSS Evening | 0 | -2 | 4 | 0 | -1 | 3 | 0.284 | 0.13 |
| VAS Mood | 2.5 | -26 | 51 | -0.5 | -15 | 18 | 0.306 | 0.36 |
| VAS Daytime Sleepiness | 0 | -47 | 25 | 4 | -10 | 17 | 0.393 | 0.01 |
| PDSS-2 Disturbed Sleep | -2 | -8 | 2 | -1 | -10 | 4 | 0.949* | NA |
| PDSS-2 Total | -2 | -8 | 2 | 0 | -10 | 4 | 0.52* | NA |

P shows p-values from robust linear mixed models (see methods section). Asterisks indicate P condition x Pre-Post. Values show averaged ratings normalized to baseline.

KSS, Karolinska Sleepiness Scale; PDSS-2, Parkinson's Disease Sleep Scale, revised version; VAS, Visual analogue scale
